# Smartphone-based App to Assess Diabetic Peripheral Neuropathy

**DOI:** 10.1101/2025.08.28.25333808

**Authors:** Rachel A. G. Adenekan, Adeyinka E. Adenekan, Kenneth K. Leung, Srikanth Muppidi, Sarada Sakamuri, Marilyn Tan, Sandra A. Tsai, Modupe Osikomaiya, Allison M. Okamura, Cara M. Nunez, Sun H. Kim, Kyle T. Yoshida

**Affiliations:** Department of Mechanical Engineering, Stanford University, 440 Escondido Mall, Building 530, Stanford, CA, 94305, USA; Independent Researcher, 108 Ilugun Rd., Abeokuta, Ogun State, Nigeria; Department of Neurology and Neurological Sciences, Stanford University, 213 Quarry Rd, Palo Alto, CA 94304, USA; Division of Endocrinology, Gerontology and Metabolism, Stanford Health Care, 300 Pasteur Drive Room S025 Stanford, CA 94305, USA; Primary Care Population Health, Stanford Health Care, 300 Pasteur Drive, Suite 305, Stanford, CA 94305, USA; School of Medicine, Stanford University, 291 Campus Drive, Stanford, CA 94305, USA; Sibley School of Mechanical and Aerospace Engineering, Upson Hall, 124 Hoy Rd, Ithaca, NY 14850, USA607; Mechanical and Aerospace Engineering, UCLA, 420 Westwood Plaza, Los Angeles, CA 90095, USA

**Author notes:** Corresponding Author: Rachel A. G. Adenekan. these authors contributed equally to this work (Cara M. Nunez, Sun H. Kim, Kyle T. Yoshida). Adeyinka E. Adenekan; Kenneth K. Leung; Srikanth Muppidi; Sarada Sakamuri; Marilyn Tan; Sandra A. Tsai; Modupe Osikomaiya; Allison Okamura; Cara M. Nunez, -255-4326; Sun H. Kim; Kyle T., Yoshida.

**Keywords:** diabetes foot exam, diabetic peripheralneuropathy, digital health, early detection, remote patient monitoring, self-monitoring

## Abstract

**Background:** Diabetic peripheral neuropathy (DPN) affects approximately 50% of individuals with diabetes and is a risk factor for amputations. Unfortunately, foot exams and screening tools are inconsistent and miss early-stage nerve damage. A smartphone-based application that delivers controlled vibrations, records patient responses, and computes a vibration perception thresh-old (SVPT) may present an accessible, precise monitoring avenue. This study assesses the clinical relevance and precision of SVPTs for measuring large-fiber sensory deficits in patients with diabetes.

**Methods:** We measured SVPTs in 71 patients with pre-diabetes or diabetes and compared their efficacy with tuning fork exams. We analyzed the correlation between SVPT and Rydel-Seiffer tuning fork (RSTF) scores, along with their relationship with clinical DPN markers such as hemoglobin A1c (HbA1c), age, and disease duration using multivariable linear regression.

**Results:** SVPTs moderately correlated with RSTF scores (*Rs* = −0.43, *p* = 0.0019). Among adults aged 50 to 69, SVPTs correlated significantly with clinical markers (*F* (4, 29) = 4.76, *p* = 0.00447, *Multiple R*^2^ = 0.396, *Adjusted R*^2^ = 0.313, *ɛ* = 0.167). The interaction between age and HbA1c was positively associated with SVPTs (*β* = 0.118, *p* = 0.001), while SVPTs were negatively associated with diabetes duration (*β* = −0.098, *p* = 0.003).

**Conclusions:** We present a clinically relevant, patient-operated smartphone application for large-fiber sensory monitoring, tested on patients with varying DPN risk. This novel platform has the potential to provide a precise, reliable, and accessible avenue for identifying individuals at risk of developing DPN complications, prior to overt clinical manifestation.

## Introduction

Diabetic peripheral neuropathy (DPN) affects approximately 50% of individuals with diabetes, resulting in pain, infections, ulcerations, amputations, sleep disturbances, reduced daily function, and higher medical costs [1, 2, 3]. While various tools exist for DPN screening and monitoring, the lack of high-resolution, accessible, fast, inexpensive, and standardized methods has led to under-diagnosis of DPN, resulting in unnecessary medical burdens [2, 4].

Common DPN screening methods, such as monofilament and tuning fork exams, are fast, inexpensive, and easy to perform, but have limited resolution and fail to detect early-stage, large-fiber nerve damage [1, 5, 6]. Studies indicate that foot screening is only done in about 50% of individuals with diabetes [7], and these tests occur less often due to increases in telehealth [8]. Precise exams, like nerve conduction studies, are inaccessible due to cost and exam duration [9]. Hence, a precise, accurate, accessible tool that monitors changes in peripheral nerve health over time could revolutionize approaches to patient care.

Smartphone vibration apps show potential to measure DPN-related vibration perception thresholds (VPTs) and have been found to discriminate between healthy patients and patients with severe DPN [10, 11]. Torres et al. characterized smart-phone vibrations and found that smartphone-based VPTs (SVPTs) correlate with monofilament thresholds on the index fingers of patients without diabetes [12]. Adenekan et al. reliably measured SVPTs in age-diverse, healthy adults, at multiple body sites, and correlated the SVPTs with tuning fork-based VPTs [13, 14]. Piaggio et al. developed a smartphone-based DPN exam using a spherical attachment to transmit smartphone vibrations to the body, used in combination with a two-point discrimination exam [15]. Most recently, Owen et al. correlated index finger SVPTs to monofilaments, tuning forks, and nerve conduction studies for healthy volunteers [16].

While these studies move toward developing an effective VPT-based DPN screening method, none of these studies provide a platform that is housed entirely in a smartphone, patient-operated, fast (*<* 2 minutes), robust, based on validated, quantitative psychophysical methods, and performs well prior to severe DPN development. Existing studies also lack analysis of how SVPTs correlate with clinical features of DPN progression like blood glucose control, age, and disease duration. Therefore, in this work we assess how SVPTs correlate with clinical markers of DPN in people with varying risk and demonstrate that SVPTs could potentially aid in earlier identification and monitoring of those patients.

## Methods

### Study Design

This was a cross-sectional study to evaluate a novel SVPT tool with traditional methods for large-fiber DPN assessment (RSTF exam and monofilament exam). This study was approved by the Stanford University Institutional Review Board (Protocol 63873). All participants provided written informed consent.

### Eligibility Criteria

Individuals aged 18 years and older and diagnosed with pre-diabetes or type-2 diabetes were recruited primarily via visits to the Stanford Endocrinology Clinic or by phone (with their treating physician’s permission). Some participants were recruited through Stanford pre-diabetes nutrition classes, campus and online flyers, and Stanford’s World Diabetes Day Fair.

We excluded patients with known touch perception deficits identified through standard monofilament exams conducted by their clinicians, as these patients have severe sensory impairments. We aimed to determine whether our tool could monitor early changes in large-fiber nerve health.

### Study Procedure and Data Collection

Participants completed a single one-hour session consisting of a low-frequency and high-frequency SVPT exam, a RSTF exam, and a monofilament exam. Exam ordering was randomized. All measurements were collected on the participant’s dominant foot.

### Medical History

Prior to completing the study, participants completed a pre-survey that inquired about demographic information, diabetes duration, presence of common neuropathy symptoms, and foot dominance. Participants also completed the Michigan Neuropathy Screening Instrument questionnaire (MNSIq [17]) to identify DPN symptoms. Anthropomorphic measures and laboratory measurements (i.e. body mass index and HbA1c) were assessed from electronic health records (EHR).

### Touch Sensitivity Measurement Methods

Figure 1 provides an overview of the setup for the SVPT exams. We used an iPhone app that modulates smartphone vibrations to stimulate a user’s foot at different intensity levels. Through this app, users respond to vibrations and the app calculates their vibration perception threshold, a recognized indicator of large-fiber nerve health [13, 14]. Specifically, this app consisted of a low and high-frequency SVPT exam on an iPhone 14 Pro Max. During the low-frequency SVPT exam, vibration frequency was held constant at 80 Hz (*hapticSharpness* = 0, Apple Core Haptics Parameter), while the amplitude (*hapticIntensity*, Apple Core Haptics Parameter), was autonomously modulated using the staircase method ([13, 14]) and visualized in Figure 1C. This method was also used for the high-frequency exam except that the vibrations were held constant at 230 Hz (*hapticSharpness* = 1). These two frequencies stimulate the Meissner and Pacinian corpuscles, which are largely responsible for vibrotactile perception and are both impaired by DPN progression [18, 19, 20, 21].

**Figure 1:**
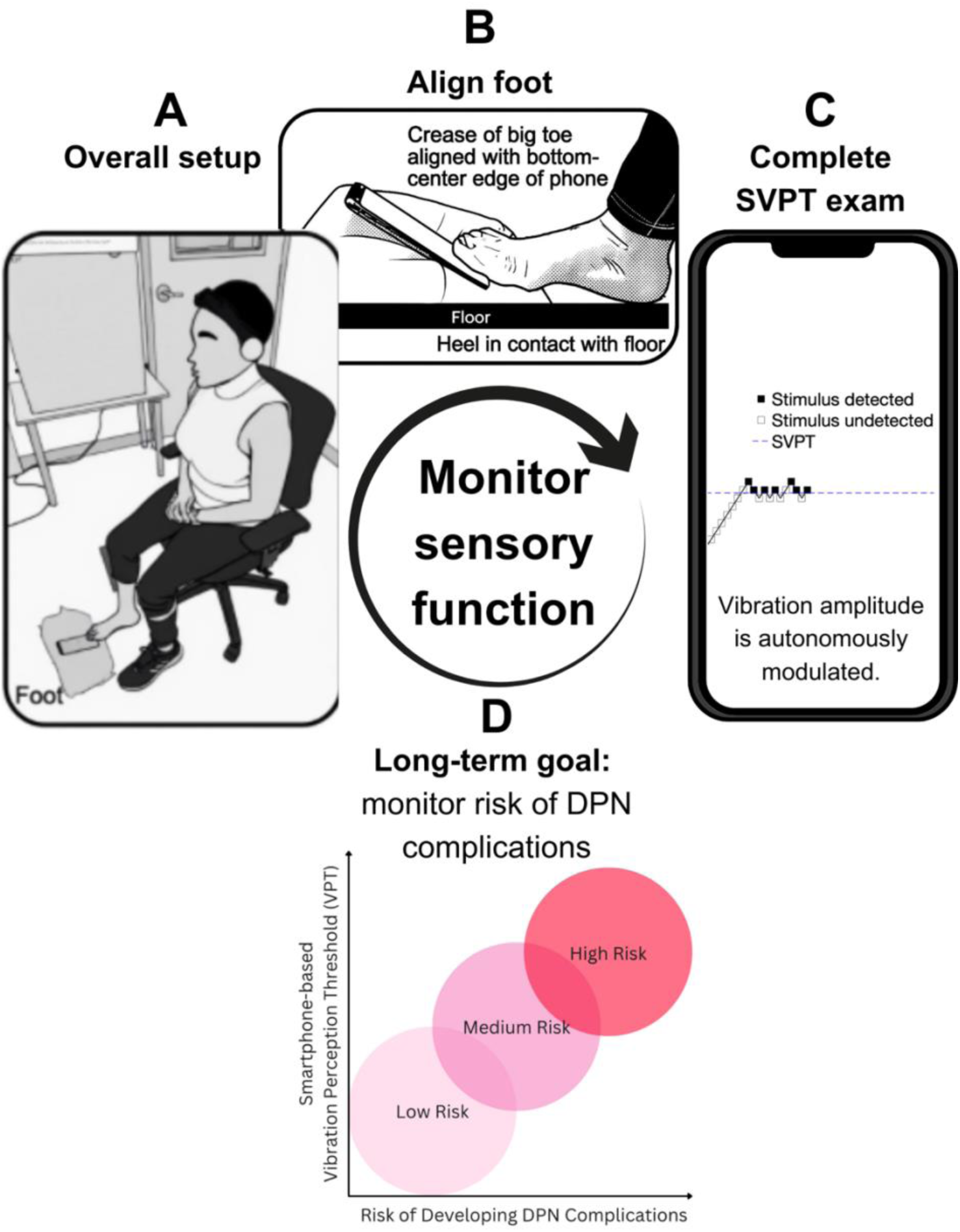
Smartphone-based vibration perception threshold (SVPT) setup and overview. (A-B) Users can complete the SVPT exam independently by placing their foot on the phone. (C) The phone then autonomously modulates the amplitude of various vibration stimuli at a given stimulation frequency. When users provide a verbal cue that they detected the vibration, the vibration amplitude is decreased. Otherwise, the vibration amplitude is increased. Upon reaching 8 reversals, the SVPT is calculated. (D) Our long-term goal is to provide an accessible platform for monitoring the risk of developing diabetic peripheral neuropathy (DPN) complications. Note: Panels (A) and (B) are adapted from Adenekan et al. 2024 [14] © IEEE 2024. Panel (C) is adapted from Adenekan et al. 2024 [13] © IEEE 2024.

Participants completed a 2-minute sample trial to familiarize themselves with the test before the measurement trials (2 minutes each). Three measurement trials were completed for each condition (high and low-frequency). Mean SVPTs for each condition are presented in the results. Participants who had SVPTs above the phone’s maximum vibration output (*hapticIntensity* = 1) are recorded as *over*, since their threshold cannot be measured with the platform. Participants wore noise-canceling headphones playing white noise to eliminate noise confounds (Figure 1A).

We used standard protocols for the 10g monofilament (Baseline Evaluation Instruments) and the 64 Hz RSTF (US Neurologicals) exams [22]. Similar to the SVPT, participants completed a sample trial and then the RSTF score was measured three times. The mean RSTF scores were analyzed.

### Statistical Analysis

#### Bivariate Analysis

To understand the relationship between SVPT and standard VPT exams, we calculated correlation coefficients between the low-frequency SVPT and RSTF score. We conducted classification analysis via area under the receiver-operating characteristic curve (ROC AUC) analysis to quantify how well SVPTs are at identifying participants with abnormal RSTF scores.

#### Multivariable Model Development

We constructed a multivariable linear model with SVPT (output) and clinical features that are associated with DPN incidence (inputs). We expected that cumulative blood glucose levels (HbA1c), diabetes duration, and age would correlate with DPN incidence based on previous research [2, 23, 24, 25]. This is our first-order model:

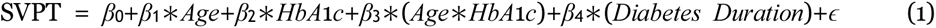

where *β*_0_ is the intercept, *β*_1_, *β*_2_, *β*_3_, and *β*_4_ are regression coefficients, and *ɛ* is the residual standard error. Coefficients were found via dataset fitting. This linear model allows us to preserve interpretability to validate our platform. Complex models like random forest approaches or neural nets would be prone to overfitting and lack interpretability. To account for the cumulative nature of DPN progression, we used average HbA1c measurements (over five years) in our model (see Supplemental Information).

Initial data processing for SVPTs was conducted in Matlab (Version 2022b) and statistical analyses were performed using R (Version 4.3.1).

#### Multivariable Model Evaluation

We evaluated our model by measuring variance in SVPT data captured by the selected features (via *Adjusted R*^2^). Features with statistically significant impacts on SVPT were identified using model coefficients and p-values (*p <* 0.05). We investigated the model’s limitations using residual analyses as presented in the Supplemental Information.

## Results

### Participant Demographics

Seventy-one adult participants with a known history of type 2 diabetes or pre-diabetes completed this study. Three participants were excluded due to their inability to complete the protocol (*n* = 1), false positives (*n* = 1), and severely ulcerated feet (*n* = 1). MNSIq scores are present for 57 out of the 68 included participants as MNSI was added to our protocol after study initiation. As shown in Table 1, the majority of our population had type 2 diabetes, with most having an HbA1c of less than 8% and an MNSIq score of less than 7. Approximately two-thirds of the participants were women.

**Table 1:** Participant Demographics and Medical History, N = 68*.

|  |  |  |
| --- | --- | --- |
| **Disease State** | Pre-diabetes | 9 (13.2%) |
|  | Type 2 diabetes | 59 (86.8%) |
| **Age [yrs]** | 18 to 49 | 9 (13.2%) |
|  | 50 to 59 | 16 (23.5%) |
|  | 60 to 69 | 31 (45.6%) |
|  | 70+ | 12 (17.6%) |
|  | Pre-diabetes: Mean (Range) | 62.6 (43,82) |
|  | Type 2 diabetes: Mean (Range) | 61.6 (38,78) |
| **Sex assigned at birth** | Male | 24 (35.3%) |
|  | Female | 44 (64.7%) |
| **Race/Ethnicity** | Native Hawaiian / Other Pacific Islander | 1 (1.5%) |
|  | Black / of African Descent | 2 (2.9%) |
|  | Hispanic / Latino | 9 (13.2%) |
|  | Asian | 26 (38.2%) |
|  | White | 30 (44.1%) |
|  | Other (Israelite) | 1 (1.5%) |
|  | Other (Middle Eastern) | 1 (1.5%) |
|  | Other (Portuguese) | 1 (1.5%) |
|  | Other (West Indian/ Caribbean) | 1 (1.5%) |
|  | Other (Indian) | 2 (2.9%) |
| **Self-reported Symptoms:**<br>**Foot pain, numbness, or**<br>**tingling** | Pain | 10 (14.7%) |
|  | Numbness | 19 (27.9%) |
|  | Tingling | 24 (35.3%) |
| **MNSIq score** | Mean ± std | 2.0 ± 2.2 |
|  | MNSIq greater than or equal to 7 | 3 (5.3%) |
| **10g Monofilament**<br>**Abnormal Sites** | Mean ± std | 0.24 ± 0.70 |
|  | Sites greater than or equal to 1 | 10 (14.7%) |
| **HbA1c (5 yr. average) [%]** | Pre-diabetes: Mean ± std | 5.9 ± 0.4 |
| | Type 2 diabetes: Mean ± std | 7.1 ± 0.9 |
|  | Type 2 diabetes: HbA1c less than 7% | 32 (47.5%) |
|  | Type 2 diabetes: HbA1c less than 8% | 54 (79.7%) |
| **Time Since Diagnosis [yrs]** | Pre-diabetes: Mean ± std | 5.1 ± 2.8 |
| | Type 2 diabetes: Mean ± std | 13.7 ± 7.7 |
| **BMI [kg/m<sup>2</sup>]** | Mean ± std | 29.6 ± 7.3 |
| **Footedness** | Right | 60 (88.2%) |
|  | Left | 3 (4.4%) |
|  | Ambipedal | 1 (1.5%) |
|  | Unknown | 4 (5.9%) |

\*Although we had 71 participants, only 68 were included in the final analysis. Three were removed due to inability to complete protocol, false positives, or severely ulcerated feet. MNSIq scores are only present for 57 out of the 68 participants as MNSIq was added to the study protocol after study initiation. Note that participants were allowed to select multiple race/ethnicity identifiers.

### Smartphone-based Vibration Perception Threshold and Rydel-Seiffer Tuning Fork Correlations

There was a statistically significant, moderate negative Spearman’s correlation (*Rs* = −0.43, *p* = 1.9 × 10^−3^) between the low-frequency SVPTs and RSTF scores (Figure 2). We also found that: (1) Participants with RSTF scores lower than 3.5 (the abnormal cutoff for ages less than 85 years [26]) tend to have low-frequency SVPTs above 0.5, indicating that low-frequency SVPTs effectively identify individuals with impaired vibration perception as classified by the RSTF (left-shaded region); and (2) Participants with low-frequency SVPTs less than 0.5 tend to have RSTF scores above 3.5, further indicating that low-frequency SVPTs identify individuals with normal vibration perception (right-shaded region).

**Figure 2:**
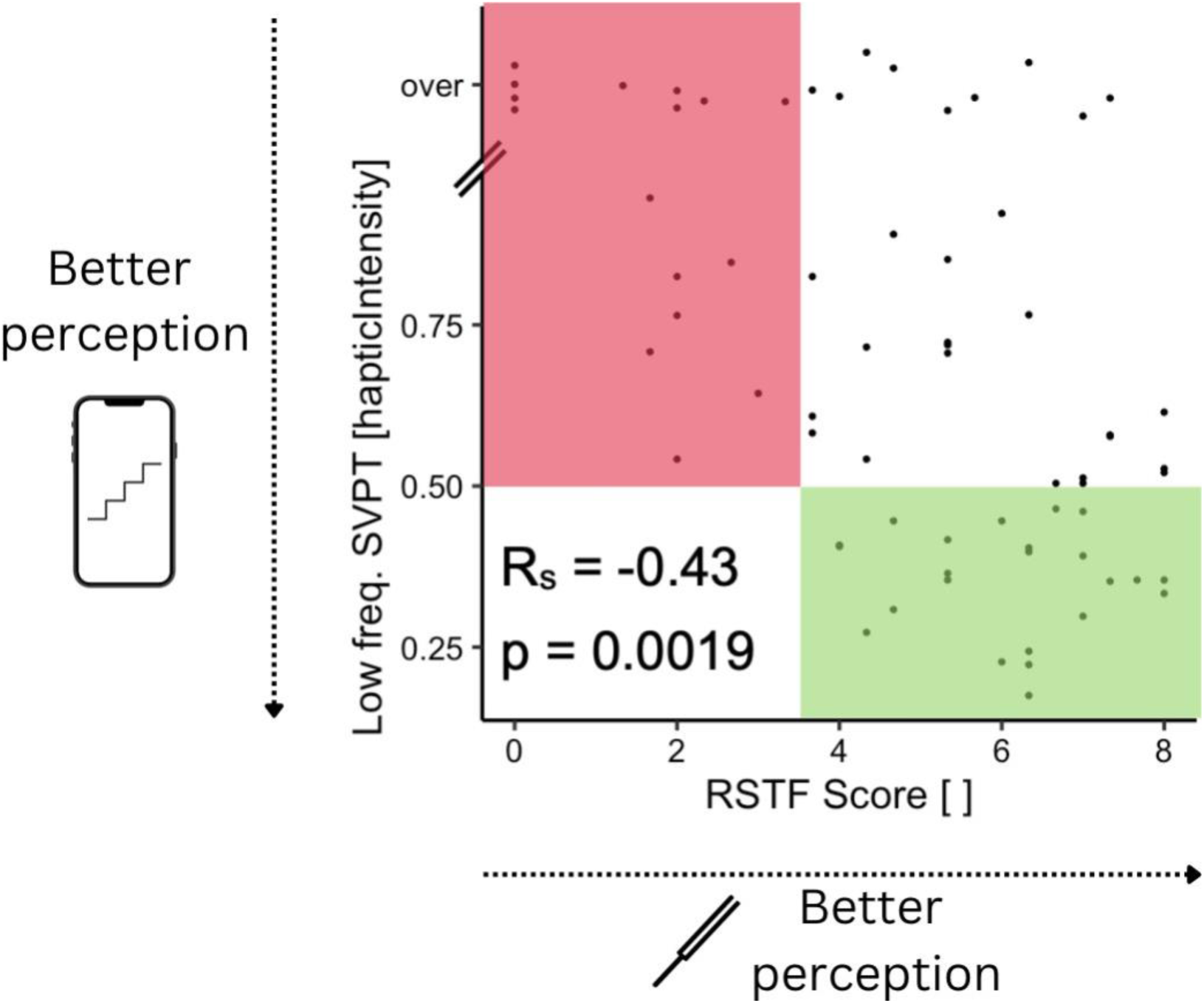
Correlations between smartphone-based vibration perception threshold (SVPT) and Rydel-Seiffer tuning fork (RSTF) score. The low-frequency SVPTs have a statistically significant, moderate correlation with the RSTF score. The smartphone-based tool can identify people with impaired vibration perception as measured on the RSTF (left-shaded region in red), and when the SVPT indicates that someone has good perception, that person also displays good perception as indicated by the RSTF score (right-shaded region in green). There is some discrepancy between the tools (detailed in the Discussion). Note: The *over* category is used for patients who have SVPTs that lie above the phone’s maximum vibration amplitude output.

Area under the receiver-operator characteristic curve (ROC AUC) analysis supported this result. Low-frequency SVPTs showed good discrimination of RSTF scores less than 3.5 from RSTF scores greater than 3.5 (*ROC AUC* = 0.86, *95% CI* = [0.75, 0.97], *p* ≤ 0.001). At the optimal cut-point of 0.54 (Youden Index), sensitivity was 100% and specificity was 67%). High-frequency SVPTs showed modest discrimination of RSTF scores (*ROC AUC* = 0.67, *95% CI* = [0.51, 0.82], *p* = 0.03). At the optimal cut-point of 0.39 (Youden Index), sensitivity was 100% and specificity was 55%).

### Ceiling and Floor Effects of Age on Smartphone-based Vibration Perception Threshold

An important requirement for the SVPT platform is its ability to discriminate between normal, age-related VPT declines [27] and DPN-related impairments. Analyzing our study’s age-diverse population, we observed that there is a statistically significant, moderate, positive Spearman’s correlation between low-frequency SVPT and age (Figure 3A, *Rs* = 0.31, *p* = 0.027) and high-frequency SVPT and age (Figure 3B, *Rs* = 0.28, *p* = 0.047). This moderate correlation is consistent with existing literature examining the effects of age on VPT using other devices [28]. We also observed that the SVPT platform may have limited utility for people younger than 50 as participants under 50 tended to have similar SVPTs (Figure 3). Similarly, the platform has limited utility for people older than 69 as these participants tended to have SVPTs above the tool’s maximum output vibration (Figure 3).

**Figure 3:**
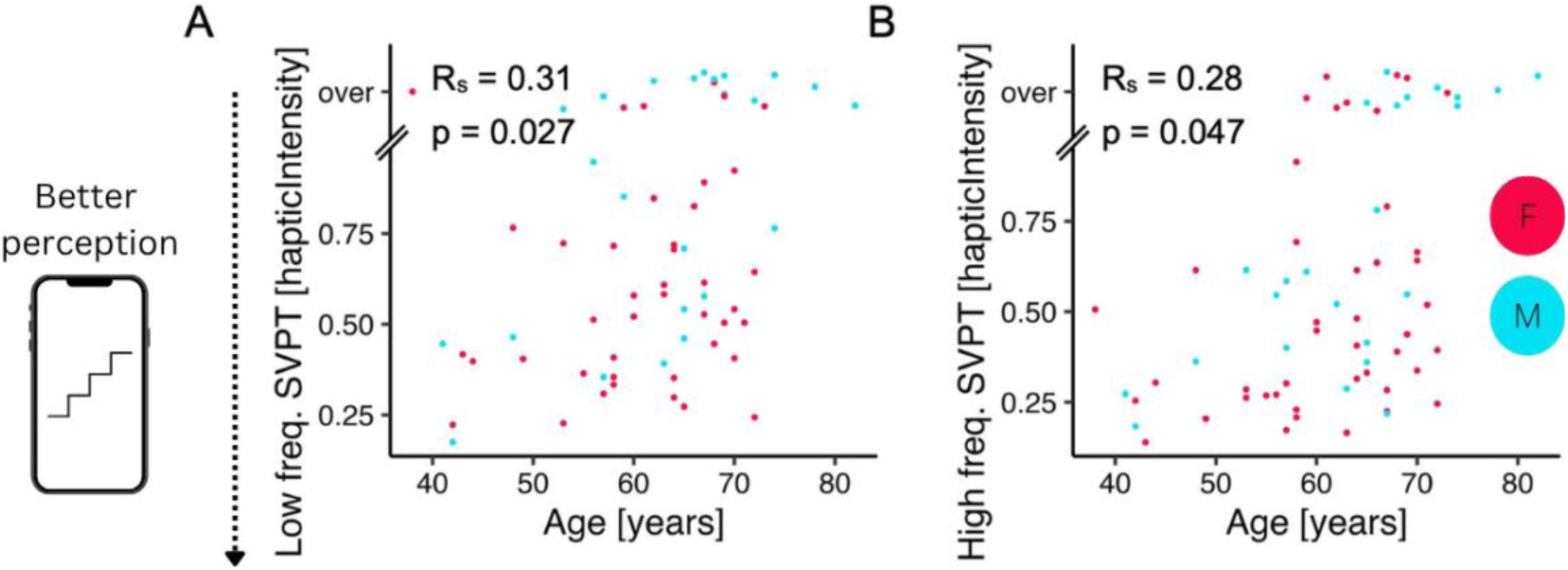
Correlations between smartphone-based vibration perception threshold (SVPT) and age. The SVPT has a statistically significant, moderate, positive correlation with age when the smartphone provides (A) low-frequency and (B) high-frequency vibrations. There are floor effects for users younger than age 50 and ceiling effects for users older than age 70. Note: The *over* category is used for patients who have SVPTs above the phone’s maximum vibration amplitude output.

Similarly, we observed a statistically significant, moderate, negative Spearman’s correlation between RSTF scores and age (Figure 4, *Rs* = −0.36, *p* = 0.002). We also observed that participants under 50 tended to have similar RSTF scores and that participants older than 75 tended to have RSTF scores that lie at the tool’s minimum output vibration.

**Figure 4:**
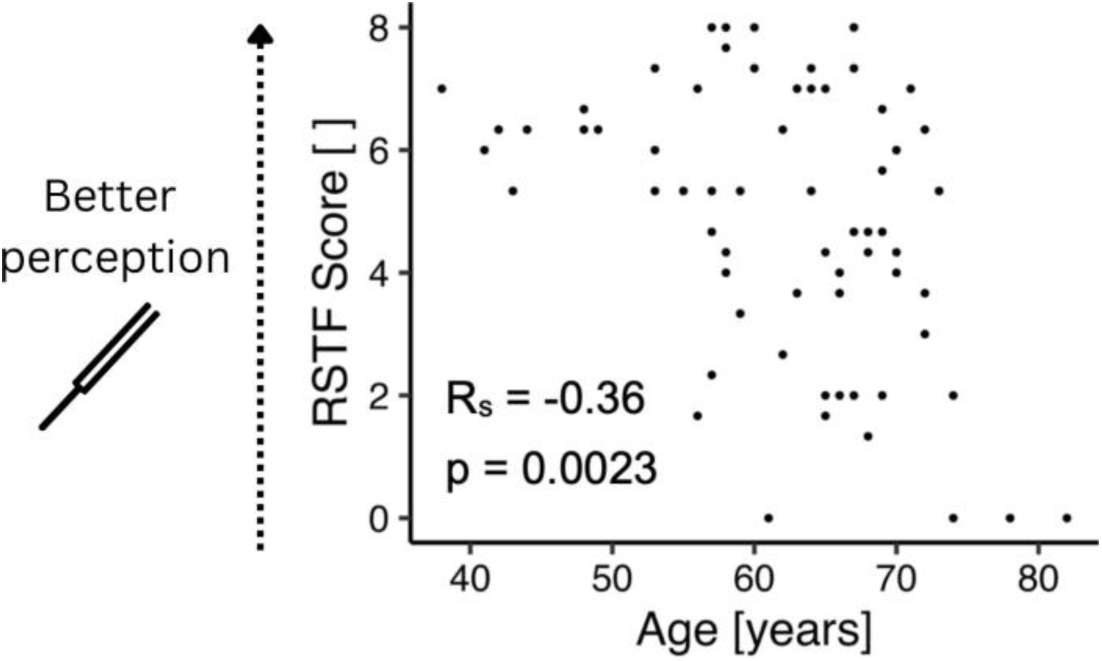
Correlations between the Rydel-Seiffer tuning fork (RSTF) score and age. There is a statistically significant, negative correlation between RSTF score and age. There are ceiling effects for patients younger than age 50 and floor effects for patients older than age 75.

### Smartphone-based Vibration Perception Threshold Prediction from a Biologically-Relevant Model

To assess the smartphone-based platform’s potential for early identification of those at risk of developing DPN complications, we constructed a multivariable regression model that predicts SVPTs from clinical features (Equation 1). Due to the previously identified floor and ceiling effects of age on SVPT, only participants who have diabetes and are aged 50 to 70 were included in the multivariable model.

For low-frequency SVPTs, the model performs moderately well. The selected features explain approximately 31% of the variance in the low-frequency SVPTs (*F* (4, 29) = 4.76, *p* = 4.47×10^−3^, *Multiple R*^2^ = 0.396, *Adjusted R*^2^ = 0.313, *ɛ* = 0.167). There was also a significant effect of diabetes duration and interaction between age and HbA1c on the SVPTs (Table 2).

**Table 2:** Biologically-Relevant Multivariable Regression Model for the Smartphone-based Vibration Perception Threshold*.

| Feature | Coefficient ($\beta$) | p-value |
| --- | --- | --- |
| Age | 0.015 | 0.604 |
| HbA1c | 0.020 | 0.505 |
| Interaction between Age and HbA1c | 0.118 | **0.001** |
| Time Since Diabetes Diagnosis | -0.098 | **0.003** |
| (Intercept) | 0.562 | $< 2 \times 10^{-16}$ |

\*Only participants who have diabetes and are between ages 50 and 70 yrs, are included in this multivariable regression model.

In contrast, a model with the same input features, but with an output of high-frequency SVPT, does not explain the variance (*F* (4, 30) = 1.03, *p* = 0.409, *Multiple R*^2^ = 0.121, *Adjusted R*^2^ = 0.003, *ɛ* = 0.189). A similar model using clinical markers to predict RSTF scores also did not fit the data (*F* (4, 29) = 1.89, *p* = 0.139, *Multiple R*^2^ = 0.207, *Adjusted R*^2^ = 0.0976, *ɛ* = 1.921).

## Discussion

Our main finding is that SVPTs provide increased precision and accuracy compared to RSTF scores and may have the potential to detect vibration perception deficits earlier than standard tools.

### Smartphone-based Vibration Perception Threshold and Rydel-Seiffer Tuning Fork Correlations

#Figure 2 and the ROC AUC analysis results show that all participants with a low, low-frequency SVPT have a RSTF score that suggests better vibration perception (right-shaded region in green). This suggests SVPTs have high sensitivity for detecting unimpaired vibratory sense. Additionally, all participants with a RSTF score that suggests impaired vibration perception have a high, low-frequency SVPT (left-shaded region in red). However, there is some disagreement between the low-frequency SVPT and the RSTF score (right-unshaded region); a high RSTF score does not guarantee that an individual will have a low, low-frequency SVPT.

This discrepancy is likely because the RSTF and the SVPT platform have different stimulation patterns. The SVPT platform uses a dynamic staircase algorithm to calculate SVPT while the RSTF uses a constantly decaying stimulus. During the staircase algorithm, vibrations start small, increase until a person feels the vibration, decrease until they do not, increase again, and so on until a set number of reversals have occurred (*n* = 8 for our current platform), at which point the SVPT is calculated by averaging the vibration amplitudes at each of the reversal points. In essence, for each SVPT measurement, there are eight data points (reversals), while the RSTF has one. During the RSTF exam, vibrations start with a high amplitude and decrease over approximately 20 seconds. Delays in a participant indicating that they no longer feel the vibration would result in a higher RSTF score, corresponding to a recorded value that is better than the participant’s true perception score. Hence the RSTF may be prone to overestimating someone’s perception ability. This matches existing literature showing that fewer reversals result in threshold overestimates and larger variances [29]. Thus we are not concerned that the low-frequency SVPTs do not perfectly correlate with the RSTF scores, as the SVPT may present a more accurate measure. Low-frequency SVPTs might be able to detect vibration perception deficits earlier than RSTF scores.

The ROC AUC analysis suggests that low-frequency SVPTs were more effective than high-frequency SVPTs in predicting abnormal RSTF scores. This is expected since RSTF’s vibration frequency of 64 Hz is closer to the low-frequency SVPT’s 80 Hz, than the high-frequency SVPT’s 230 Hz. Still, it is encouraging that both low- and high-frequency SVPTs may be useful in identifying sensory deficits.

### Ceiling and Floor Effects of Age on Smartphone-based Vibration Perception Threshold

The results in Figures 3 and 4 show that participants younger than 50 had low, low-frequency SVPTs indicating better vibratory perception. DPN is a progressive condition that develops over time due to elevated blood glucose levels and other metabolic dysfunction [2]. Individuals under 50 may be more resilient to the effects of hyperglycemia than their older counterparts. Conversely, adults over age 69 experience a rapid age-related decline in vibration perception threshold (VPT) [27]. Thus it is not surprising that both the SVPT platform (Figure 3) and the RSTF (Figure 4) experienced saturation in measuring VPT of people who are over age 69.

### Smartphone-based Vibration Perception Threshold Prediction from a Biologically Relevant Model

The interaction between age and HbA1c had a coefficient of *β*_3_ = 0.118 (*p* = 0.001). This suggests that younger adults maintain their SVPTs even with elevated blood glucose levels, but as patients age, elevated blood glucose levels are associated with higher SVPTs (impaired vibrotactile perception) as expected [30]. However, diabetes duration had a coefficient of *β*_4_ = −0.098 (*p* = 0.003), suggesting that longer diabetes duration is associated with lower SVPTs (better VPT), contrary to expectation. This result might stem from our exclusion of patients with severe neuropathy. Most of our patients had an MNSIq score of less than 7, and we excluded participants with ulcerated feet or known inability to feel the 10g monofilament in the pad of the big toe from the study. As such, patients with the worst DPN cases, with long-term metabolic dysfunction, are not included in our study.

Additionally, most study participants were being treated for diabetes in the Stanford Endocrinology Clinic. Our unexpected result (higher diabetes duration corresponding to improved vibrotactile perception) could be a function of health equity. It might be that these patients have had better access to early endocrinology care, yielding better diabetes management and improved SVPTs. Given that many people are unaware of pre-diabetes [2, 31, 32], diabetes duration, recorded as self-reported time since diagnosis, does not exactly translate to time since initial sustained presentation of elevated blood glucose levels. Diabetes duration is more likely a measure of how long someone has had access to care than of exactly how long someone has had elevated blood glucose levels. We do not have the dataset required to thoroughly assess this hypothesis.

Still, the model’s moderate predictive power of low-frequency SVPT from clinical features suggests that SVPTs have the potential to aid in earlier identification of those at risk of developing DPN complications. Unlike the low-frequency SVPTs, high-frequency SVPTs did not correlate with clinical markers of DPN incidence in this study. Current literature suggests that both Meissner (sensitive to low frequencies) and Pacininan corpuscles (sensitive to high frequencies) are affected by DPN, but because they may be differentially impaired [20, 21, 33], the low- and high-frequency SVPTs may yield different findings.

### Limitations

While this study yielded promising insights, there are limitations. First, this study had a small sample size (*n* = 71) and was conducted at a single site. Nevertheless, our patients had HbA1c levels and symptoms that were representative of the broader population of Americans living with diabetes ([34] and Table 1). Another limitation is that SVPT primarily assesses large fiber nerve damage and cannot detect small-fiber neuropathy. This limitation also applies to current screening methods including the monofilament and tuning fork exams, as well as the more precise, but invasive, electromyography-nerve conduction studies (EMG-NCS). Additionally, we did not compare the SVPT exam to the gold standard for quantifying large-fiber nerve damage, EMG-NCS, because most of our patients did not have an indication for EMG-NCS. SVPTs, like other diagnostic tools (EMG-NCS, small fiber testing, etc.), are not intended to be interpreted in isolation, but rather to provide an additional data point for a trained physician to understand the patient’s broader clinical context. A strength of using the RSTF in this study is that it provides a standard method of application and quantitative measurements. The inter- and intra-rater reliability and responsiveness (sensitivity to change) in monitoring patients with varying degrees of polyneuropathy (normal, mild, moderate, severe) has been demonstrated in several studies ([26, 35, 36]), and comparing the RSTF with the SVPT allows validation of SVPT as one tool in the diagnosis of neuropathy focused on large-fiber function.

Despite some limitations, this study sets the foundation for digital technology that could be used to monitor large-fiber sensory function in adults with diabetes. In clinical practice, many patients with a diagnosis of pre-diabetes or diabetes may have no self-reported, overt symptoms of neuropathy, but are found on neurologic examination testing to have sensory deficits indicative of neuropathy. SVPTs may be helpful in this regard and may have the additional benefit of being patient-operated, housed entirely within a smartphone, and easily performed outside of the physician’s office.

## Conclusions

We assessed the validity of a smartphone-based vibration perception threshold (SVPT) compared to traditional, clinical measures of large-fiber sensory function, in people with pre-diabetes and diabetes. We found that SVPTs moderately correlate with Rydel-Seiffer tuning fork-based vibration perception thresholds and that SVPTs of patients with diabetes aged 50 to 69 moderately correlate with indicators of DPN incidence (age, blood glucose control, and time since diagnosis).

To our knowledge, we present the first clinically relevant platform for large-fiber sensory monitoring tested on patients with varying DPN risk that is fast, patient-operated, and housed entirely within a smartphone. This platform has the potential to provide an accessible avenue for identifying individuals at risk of developing DPN complications, prior to overt clinical manifestation. Large, longitudinal, and prospective SVPT datasets paired with more sophisticated and inclusive models are needed to determine whether SVPT can enhance prediction of DPN and its associated complications in diverse populations. The diagnosis of DPN must be made in light of the overall clinical presentation and diagnostic testing, under the guidance of a trained physician. SVPTs may potentially be used, in combination with other diagnostic tools, to enable earlier understanding of a patient’s overall clinical presentation.

## Data Availability

All data produced in the present study are available upon reasonable request to the authors.

## Abbreviations

DPN: diabetic peripheral neuropathy
SVPT: smartphone-based vibration perception threshold
RSTF: Rydel-Seiffer tuning fork
HbA1c: hemoglobin A1c
VPT: vibration perception threshold
ROC AUC: area under the receiver-operating characteristic curve
MNSIq: Michigan Neuropathy Screening Instrument questionnaire
EHR: electronic health record
BMI: body mass index
CI: confidence intervals
EMG-NCS: electromyography-nerve conduction study

## Funding Source

This work was supported in partby the Precision Health and Integrated Diagnostics Center at Stanford, the Stanford Center for Digital Health, the Stanford Diabetes Research Center (National Institutes of Health award no. P30DK116074), the National Science Foundation Graduate Research Fellowship Program, the Stanford Graduate Fellowship Program, and the Stanford Summer Undergraduate Research Fellowship Program.

## Conflict of Interest Disclosure

Not applicable

## Acknowledgements

We thank all the study participants and clinic staff.

## Supplemental Information

### HbA1c Measurements

To account for the cumulative nature of DPN progression, we used average HbA1c measurements (over five years) in our model. Over the five-year period, the median and interquartile range of the HbA1c entry count per patient were 10 and 8.25 respectively. Measurements over this period were typically available within the Stanford EHR of many patients; longer periods, such as 10 years, were not available for many patients. There has also been some recent literature suggesting that HbA1c variability, in addition to mean, is correlated with DPN progression [25]. We did not use HbA1c variability for this study because the data was sparse (many patients lacked regular readings for the entire 5-year period).

### Regression Model Limitations

To assess the limitations of our multivariable regression model, we conducted residual analysis (Figure S1). From these figures, we observed that although the variance of the residuals is not perfectly consistent for all observations, there is no obvious pattern and thus no cause for concern (Figure S1A). From Figure S1B, we observed that there are no extreme outliers in our dataset (points that lie outside of Cook’s distance), but there are some points that are close to having extreme influence (points near Cook’s distance line). Lastly, the residuals of our regression model can be assumed to be normally distributed as evidenced by the points in our Quantile-Quantile residuals plot falling roughly along a straight diagonal line (Figure S1C).

---

**Figure S1:**
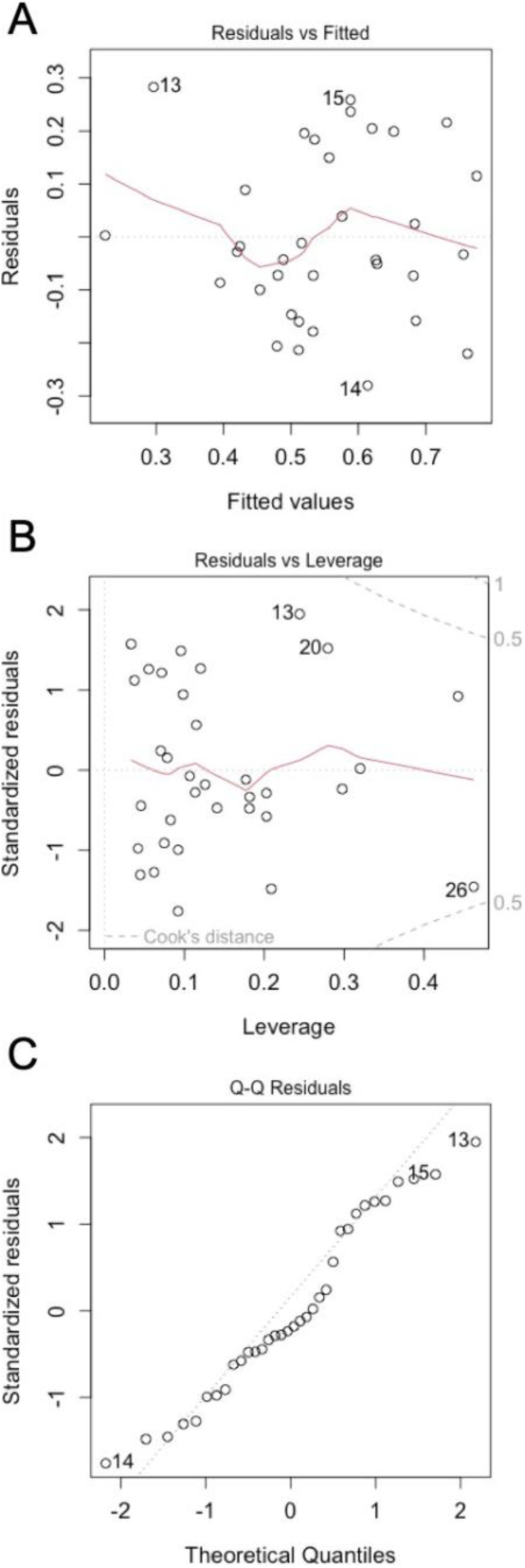
Residual analysis of the model. (A) Plot of the residuals vs. the fitted values. Although the variance of the residuals is not perfectly consistent for all observations, there is no obvious pattern and thus no cause for concern. (B) Plot of the standardized residuals vs. the leverage. There are no extreme outliers in our dataset (points that lie outside of Cook’s distance), but there are some points that are close to having extreme influence. (C) A Quantile-Quantile (Q-Q) plot of the standardized residuals vs. the theoretical quantiles. The points in this plot fall roughly along a straight diagonal line which suggests normally distributed residuals in the regression model.

## Notes

### Competing Interest Statement

The authors have declared no competing interest.

### Funding Statement

This work was supported in part by the Precision Health and Integrated Diagnostics Center at Stanford, the Stanford Center for Digital Health, the Stanford Diabetes Research Center (National Institutes of Health award no. P30DK116074), the National Science Foundation Graduate Research Fellowship Program, the Stanford Graduate Fellowship Program, and the Stanford Summer Undergraduate Research Fellowship Program.

### Author Declarations

Institutional Review Board of Stanford University gave ethical approval for this work.

### Summary of Updates

We made significant improvements to the paper including adding additional data to Table 1, including ROC AUC analysis, and revising the discussion.

## References

[1] Pop-Busui R, Boulton AJ, Feldman EL, Bril V, Freeman R, Malik RA, Sosenko JM, Ziegler D. Diabetic Neuropathy: A Position Statement by the American Diabetes Association. Diabetes Care. 2017;40:136–54.

[2] Pop-Busui R, Ang L, Boulton AJ, Feldman EL, Marcus RL, Mizokami-Stout K, Singleton JR, Ziegler D. Diagnosis and treatment of painful diabetic peripheral neuropathy. ADA Clinical Compendia. 2022;1:1–32.

[3] Gordois A, Scuffham P, Shearer A, Oglesby A, Tobian JA. The health care costs of diabetic peripheral neuropathy in the US. Diabetes Care. 2003;26(6):1790–5.

[4] Sloan G, Selvarajah D, Tesfaye S. Pathogenesis, diagnosis and clinical management of diabetic sensorimotor peripheral neuropathy. Nature Reviews Endocrinology. 2021;17(7):400–20.

[5] Rossing TD, Russell DA, Brown DE. On the acoustics of tuning forks. American Journal of Physics. 1992;60(7):620–6.

[6] Dros J, Wewerinke A, Bindels PJ, van Weert HC. Accuracy of Monofilament Testing to Diagnose Peripheral Neuropathy: A Systematic Review. Annals of Family Medicine. 2009;7(6):555–8.

[7] Patel J, Zamzam A, Syed M, Blanchette V, Cross K, Albalawi Z, Al-Omran M, De Mestral C. A scoping review of foot screening in adults with diabetes mellitus across Canada. Canadian Journal of Diabetes. 2022;46(5):435–40.

[8] Arao KA, Fincke BG, Zupa MF, Vimalananda VG. Comparison of Endocrinologists’ Physical Examination Documentation for In-person vs Video Telehealth Diabetes Visits. Journal of the Endocrine Society. 2023;7(7):1–5. Bvad073.

[9] Zhao Z, Ji L, Zheng L, Yang L, Yuan H, Chen L, Shan Z, Shen S, Li Q, Shi B, et al. Effectiveness of clinical alternatives to nerve conduction studies for screening for diabetic distal symmetrical polyneuropathy: A multi-center study. Diabetes research and clinical practice. 2016;115:150–6.

[10] May JD, Morris MWJ. Mobile phone generated vibrations used to detect diabetic peripheral neuropathy. Foot and Ankle Surgery. 2017;23(4):281–4.

[11] Jasmin M, Yusuf S, Syahrul S, Abrar EA. Validity and Reliability of a Vibration-Based Cell Phone in Detecting Peripheral Neuropathy among Patients with a Risk of Diabetic Foot Ulcer. The International Journal of Lower Extremity Wounds. 2021;22(4):687–94.

[12] Torres WO, Abbott ME, Wang Y, Stuart HS. Skin Sensitivity Assessment Using Smartphone Haptic Feedback. IEEE Open Journal of Engineering in Medicine and Biology. 2023;4:216–21.

[13] Adenekan RAG, Reyes AG, Yoshida KT, Kodali S, Okamura AM, Nunez CM. A Comparative Analysis of Smartphone and Standard Tools for Touch Perception Assessment Across Multiple Body Sites. IEEE Trans-actions on Haptics. 2024;17(4):970–7. Doi:10.1109/TOH.2024.3362154.

[14] Adenekan RAG, Yoshida KT, Benyoucef A, Reyes AG, Adenekan AE, Okamura AM, Nunez CM. Reliability of Smartphone-Based Vibration Threshold Measurements. In: IEEE Haptics Symposium; 2024. p. 25–32.

[15] Piaggio D, Castaldo R, Garibizzo G, Iadanza E, Pecchia L. A smartphone-based tool for screening diabetic neuropathies: A mHealth and 3D printing approach. Biomedical Signal Processing and Control. 2024;89:1–10. 105807.

[16] Lindsay OR, Hammad H, Baysic J, Young A, Osman N, Ferber R, Culos-Reed N, Peters RM. Age related changes in skin sensitivity assessed with smartphone vibration testing. Scientific Reports. 2024;14(1):1–12.

[17] Feldman EL, Stevens M, Thomas P, Brown M, Canal N, Greene D. A practical two-step quantitative clinical and electrophysiological assessment for the diagnosis and staging of diabetic neuropathy. Diabetes Care. 1994;17(11):1281–9.

[18] Verrillo RT. Vibration sensation in humans. Music Perception. 1992;9(3):281–302.

[19] Johansson RS, Flanagan JR. Coding and use of tactile signals from the fingertips in object manipulation tasks. Nature Reviews Neuroscience. 2009;10(5):345–59.

[20] García-Mesa Y, Feito J, González-Gay M, Martínez I, García-Piqueras J, Martín-Cruces J, Viña E, Cobo T, García-Suárez O. Involvement of cutaneous sensory corpuscles in non-painful and painful diabetic neuropathy. Journal of Clinical Medicine. 2021;10(19):4609.

[21] Vega JA, García-Suárez O, Montaño JA, Pardo B, Cobo JM. The Meissner and Pacinian sensory corpuscles revisited new data from the last decade. Microscopy Research and Technique. 2009;72(4):299–309.

[22] Panosyan FB, Mountain JM, Reilly MM, Shy ME, Herrmann DN. Rydel-Seiffer fork revisited: Beyond a simple case of black and white. Neurology. 2016;87(7):738–40.

[23] Lian X, Qi J, Yuan M, Li X, Wang M, Li G, Yang T, Zhong J. Study on risk factors of diabetic peripheral neuropathy and establishment of a prediction model by machine learning. BMC Medical Informatics and Decision Making. 2023;23(1):146.

[24] Casadei G, Filippini M, Brognara L. Glycated hemoglobin (HbA1c) as a biomarker for diabetic foot peripheral neuropathy. Diseases. 2021;9(16):1–18.

[25] Tesfaye S, Stevens L, Stephenson J, Fuller J, Plater M, Ionescu-Tirgoviste C, Nuber A, Pozza G, Ward J, Group EICS. Prevalence of diabetic peripheral neuropathy and its relation to glycaemic control and potential risk factors: the EURODIAB IDDM Complications Study. Diabetologia. 1996;39:1377–84.

[26] Martina IS, van Koningsveld R, Schmitz PI, van der Meché FG, van Doorn PA. Measuring vibration threshold with a graduated tuning fork in normal aging and in patients with polyneuropathy. Journal of Neurology, Neurosurgery & Psychiatry. 1998;65(5):743–7.

[27] Halonen P. Quantitative vibration perception thresholds in healthy subjects of working age. European Journal of Applied Physiology and Occupational Physiology. 1986;54:647–55.

[28] Wells C, Ward LM, Chua R, Inglis JT. Regional variation and changes with ageing in vibrotactile sensitivity in the human footsole. Journal of Gerontology: Biological Sciences. 2003;58A(8):680–6.

[29] Witton C, Talcott JB, Henning GB. Psychophysical measurements in children: challenges, pitfalls, and considerations. PeerJ. 2017;5:1–22.

[30] Pfannkuche A, Alhajjar A, Ming A, Walter I, Piehler C, Mertens PR. Prevalence and risk factors of diabetic peripheral neuropathy in a diabetics cohort: Register initiative “diabetes and nerves”. Endocrine and Metabolic Science. 2020;1(1-2):1–9.

[31] Sun H, Saeedi P, Karuranga S, Pinkepank M, Ogurtsova K, Duncan BB, Stein C, Basit A, Chan JC, Mbanya JC, et al. International Diabetes Federation Diabetes Atlas: Global, regional and country-level diabetes prevalence estimates for 2021 and projections for 2045. Diabetes Research and Clinical Practice. 2022;183:109119.

[32] Prevention C. National diabetes statistics report. Atlanta, GA: Centers for Disease Control and Prevention, US Department of Health and Human Services. 2020. Available from: https://www.cdc.gov/diabetes/php/data-research/index.html. Accessed 2024 May 19.

[33] Lindholm E, Löndahl M, Fagher K, Apelqvist J, Dahlin LB. Strong as-sociation between vibration perception thresholds at low frequencies (4 and 8 Hz), neuropathic symptoms and diabetic foot ulcers. PLOS ONE. 2019;14(2):1–15. E0212921.

[34] Fang M, Wang D, Coresh J, Selvin E. Trends in diabetes treatment and control in US adults, 1999–2018. New England Journal of Medicine. 2021;384(23):2219–28.

[35] Merkies IS, Schmitz PI, van der Meché FG, van Doorn PA. Reliability and responsiveness of a graduated tuning fork in immune mediate polyneuropathies. Journal of Neurology, Neurosurgery & Psychiatry. 2000;68(5):669–71.

[36] Pestronk A, Florence J, Levine T, Al-Lozi MT, Lopate G, Miller T, Ramneantu I, Waheed W, Stambuk M. Sensory exam with a quantitative tuning fork: rapid, sensitive and predictive of SNAP amplitude. Neurology. 2004;62(3):461–4.

